# Knowledge, Attitude and Practice towards COVID-19 among people in Bangladesh during the pandemic: a cross-sectional study

**DOI:** 10.1101/2020.09.22.20198275

**Authors:** Md. Golam Rabbani, Orin Akter, Md. Zahid Hasan, Nandeeta Samad, Shehrin Shaila Mahmood, Taufique Joarder

## Abstract

The world is grappling with Covid-19, a dire public health crisis. Preventive and control measures are adopted to reduce the spread of COVID-19. It is important to know the knowledge, attitude, and practice (KAP) of people towards this pandemic to suggest appropriate coping strategies. The aim of this study was to assess the KAP of Bangladeshi people towards Covid-19 and determinants of those KAPs. We conducted a cross-sectional survey of 492 Bangladeshi people aged above 18 years from May 7 to 29, 2020 throughout the country. Simple and multiple logistic regression analyses were conducted to identify the factors associated with KAP on COVID-19. About 45% of respondents had good knowledge, 49% of respondents expressed positive attitude towards controlling of COVID-19 and 24% of respondents had favorable practice towards COVID-19. Almost three fourths of the respondents went outside home during the lockdown period. Furthermore, the study found that good knowledge and attitude were associated with better practice of COVID-19 health measures. An evidence informed and context specific risk communication and community engagement, and a social and behavior change communication strategy against COVID-19 should be developed in Bangladesh, based on the findings of this study, targeting different socio-economic groups.

## Introduction

The world is struggling with COVID-19 pandemic for quite some time, and Bangladesh is hard hit [1]. As of July 30, 2020, it has been reported across 215 countries and regions due to human interaction, and has infected more than 17 million people with 672,364 deaths [2]. It is concerning that, in terms of daily identified case rates, Bangladesh--a lower middle income country (LMIC)--has ranked 16th in the world and 3rd among the South Asian countries[2]. Although Bangladesh detected the first case later than many countries (8 March 2020), to date (July 30, 2020), a total of 234,889 cases have been identified, including total 3,083 deaths [3].

One of the reasons for such a rapid increase may be that Bangladesh is the second most densely populated country in the world [4]. Recent statistics have estimated that population of Bangladesh is about 165 million with 1,239.6 people per square kilometers [5,6]. In Bangladesh, a large proportion of population still lives below the poverty line, and almost half of the population is exposed to multiple socioeconomic vulnerabilities[4,7]. Evidently, newer underprivileged communities are falling a victim to COVID-19 [8]. Bangladesh suffers from a low literacy rate [5], which may potentially expose the population to an unfavorable knowledge, attitude and practice (KAP) towards a persisting pandemic. Socio-economic status, over population, lifestyle, etc. may contribute to the recent rapid increase of Covid-19 cases in Bangladesh.

There is no control measure and treatment considered effective to combat the pandemic except for Convalescent Plasma Therapy (CPT), until vaccine is available [9,10]. However, regarding prevention of the spread of this disease, non-clinical interventions based on primary health care practice have been suggested by the World Health Organization (WHO) considering existing scientific evidences [11]. These interventions have been proposed as the cheapest, easiest, and the most effective ways to interrupt the spread of the virus [12,13,14], but these are largely dependent on people’s KAP. Appropriate maintenance of these interventions is important to reduce the spread of outbreaks and a responsive health system can play a key role to implement social and behavior change communication (SBCC) interventions to control such outbreaks [15,16]. Public behavior is also crucial in combating the pandemic influenced by people’s knowledge of preventing this infectious disease. Recent scientific evidences have demonstrated that the adequate knowledge, attitude and appropriate practice of the interventions are associated with reduction of morbidity and mortality and ultimately total control over COVID-19 [17, 18]. Thus, coordination of whole-society in an appropriate way for generating knowledge and maintaining proper attitude and practice is essential to counter the pandemic [16]. Although it is believed that knowledge and practice measures are the ultimate solutions, the interventions such as social distancing, hand hygiene, home quarantine, etc. may seem to the people of Bangladesh as new concepts which should be ingrained. Hence, In Bangladesh, it would be difficult to get used to with the interventions in a short of time, without a thoroughly designed context specific SBCC strategy.

As of now, there have been no alternative to generating awareness against COVID-19 among the people and construct relevant KAP among them. Therefore, to facilitate management against COVID-19 in Bangladesh, it is important to understand the public’s KAP of COVID-19 and undertake necessary strategies. Although several studies related to KAP towards COVID-19 have been conducted globally, there is paucity of such study in Bangladesh that includes all divisions and conducts survey through audio communication instead of online survey. This evidence should be useful for policymakers, as it will allow them to design a context-specific social and behavior change strategy in Bangladesh. The objective of this study was to assess the KAP towards COVID-19 of Bangladeshi residents during the rapid rise period of the COVID-19 outbreak in Bangladesh.

## Materials and Methods

### Study design and setting

A cross-sectional survey from May 7 to May 29, 2020, during the lockdown period in Bangladesh was conducted among 492 individuals aged 18 years and above for measuring the KAPs regarding COVID-19. The study was conducted throughout the country as it surveyed individuals from eight administrative divisions (Barishal, Chattogram, Dhaka, Khulna, Mymensingh, Rajshahi, Rangpur, and Sylhet) of Bangladesh. As it was not feasible to conduct a community-based national sampling survey during the national lockdown or restricted-mobility period, we decided to collect the data through direct phone calls or through digital social media platforms like WhatsApp, Messenger, Skype, Zoom etc. However, we also conducted face-to-face surveys in some cases for the convenience of the respondents. Primarily, we collected contact numbers from the network of the studied population. Later, we contacted the individuals and surveyed them if consented. Additionally, from eight divisions, we hired volunteers who assisted in data collection from their respective divisions.

### Sample design

We calculated the sample size from an unknown population by using simple random sampling technique at 95% confidence interval and at 0.5 level of precision, and the sample size was determined as 384. We anticipated that around 20% of participants would not participate in the survey. Therefore, the sample size was increased to 480 after adjusting for the 20% non-response rate. We spilt the sample into eight divisions proportionately to the population of the respective divisions (Barishal 5.7%, Chattogram 17.5%, Dhaka 23.3%, Khulna 11.9%, Mymensingh 7.4%, Rajshahi 14.3%, Rangpur 11.8%, and Sylhet 6%) [19].

### Data collection instruments and measures

We developed a structured questionnaire for the individual survey which consisted of two segments: 1) Socio-demographic characteristics, and 2) Knowledge, attitude and practice. Socio-demographic variables included age, gender, education, occupation, current residence, religion, marital status, number of persons, room, toilets in current living residence and income. According to national guidelines for clinical and community management of COVID-19 by the Government of Bangladesh, WHO reports, and rigorous literature review, the investigators pilot tested a COVID-19 questionnaire [1,10,11,17]. The questionnaire includes a few questions regarding clinical presentations, transmission routes, prevention and control, and source of knowledge of COVID-19. These questions were answered on a yes or no basis with an additional “don’t know” option. A correct answer was assigned 1 point and an incorrect and don’t know answer was assigned 0 point. The total knowledge score ranged from 0 to 14, with a higher score denoting a better knowledge of COVID-19. To determine the KAP level, the cut off value was determined by authors based on the context of Bangladesh considering the ghastliness of COVID-19. Having more than 80% scores was classified as “Good knowledge” and having less than or equal to 80% scores was considered as “poor knowledge”. Similar scoring approach was used for classifying “positive attitude” and “negative attitude”, “good practice” and “poor practice”.

### Statistical Analysis

Both descriptive and inferential statistical analyses were performed. In the descriptive analyses, the characteristics of the study participants were presented in terms of frequency (n) and percentages (%) with 95% confidence interval (CI). KAPs of different groups according to demographic and socio-economic characteristics were compared. Simple logistic and multiple logistic regression analyses were conducted using all of the socio-demographic variables as exposures and knowledge as the outcome variable to identify factors associated with knowledge. Similar analyses were performed to identify factors associated with attitudes and practices. We analyzed the data using Stata version 13 and Microsoft Excel.

### Ethical approval

The study protocol was approved by Ethical Review Committee of Public Health Foundation, Bangladesh [Ethics Reference No:2020/01]. We adhered to all ethical principles during the research process.

## Results

### Demographic and socioeconomic characteristics

As shown in Table 1. Demographic and socioeconomic characteristics of participants (N = 492)., a total of 492 individuals were surveyed in this study and majority of them belonged to younger age group (52% below 35 years). Among the participants, about 65% were male, 32% had a higher level of education (bachelor or higher level), 62% were currently living in the rural area, 41% were not employed and had no income, and more than 90% of respondents had access to available running water. Other characteristics are shown in Table 1. Demographic and socioeconomic characteristics of participants (N = 492).

**Table 1.** Demographic and socioeconomic characteristics of participants (N = 492).

| Variables | n | % | 95% CI |
| --- | --- | --- | --- |
| **Age group** |  |  |  |
| ≤ 25 | 84 | 17.07 | (14-20.7) |
| 26-35 | 172 | 34.96 | (30.9-39.3) |
| 36-45 | 83 | 16.87 | (13.8-20.5) |
| 46-55 | 59 | 11.99 | (9.4-15.2) |
| 56-65 | 54 | 10.98 | (8.5-14.1) |
| ≥ 66 | 40 | 8.13 | (6-10.9) |
| **Sex** |  |  |  |
| Male | 321 | 65.24 | (60.9-69.3) |
| Female | 171 | 34.76 | (30.7-39.1) |
| **Education Level** |  |  |  |
| No education | 76 | 15.45 | (12.5-18.9) |
| Primary and can sign | 87 | 17.68 | (14.5-21.3) |
| Secondary and SSC | 103 | 20.93 | (17.6-24.8) |
| HSC passed or equivalent | 70 | 14.23 | (11.4-17.6) |
| Higher | 156 | 31.71 | (27.7-36) |
| **Occupation** |  |  |  |
| Currently not employed | 202 | 41.06 | (36.8-45.5) |
| Service holder | 134 | 27.24 | (23.5-31.4) |
| Farmer | 28 | 5.69 | (4-8.1) |
| Business man | 57 | 11.59 | (9-14.7) |
| Day labor | 52 | 10.57 | (8.1-13.6) |
| Others | 19 | 3.86 | (2.5-6) |
| **Religion** |  |  |  |
| Muslim | 454 | 92.28 | (89.6-94.3) |
| Others | 38 | 7.72 | (5.7-10.4) |
| **Current living residence** |  |  |  |
| Urban | 188 | 38.21 | (34-42.6) |
| Rural | 304 | 61.79 | (57.4-66) |
| **Marital status** |  |  |  |
| In a marital relationship | 331 | 67.28 | (63-71.3) |
| Not in a marital relationship | 161 | 32.72 | (28.7-37) |
| **Family size** |  |  |  |
| 1-3 members | 109 | 22.15 | (18.7-26.1) |
| 3-6 members | 316 | 64.23 | (59.9-68.4) |
| 7 & more | 67 | 13.62 | (10.9-17) |
| **Earning person in family** |  |  |  |
| No earning person | 6 | 1.22 | (0.5-2.7) |
| Single earning person | 253 | 51.42 | (47-55.8) |
| Two & more | 233 | 47.36 | (43-51.8) |
| **Monthly income** |  |  |  |
| No earning | 202 | 41.06 | (36.8-45.5) |
| 1000-10000 | 111 | 22.56 | (19.1-26.5) |
| 11000-20000 | 80 | 16.26 | (13.2-19.8) |
| 21000-30000 | 39 | 7.93 | (5.8-10.7) |
| 31000-40000 | 33 | 6.71 | (4.8-9.3) |
| >40000 | 27 | 5.49 | (3.8-7.9) |
| **Availability of running water at home** |  |  |  |
| Yes | 449 | 91.26 | (88.4-93.5) |
| No | 43 | 8.74 | (6.5-11.6) |

| **Division** |  |  |  |
| --- | --- | --- | --- |
| Barishal | 30 | 6.1 | (4.3-8.6) |
| Chattogram | 87 | 17.68 | (14.5-21.3) |
| Dhaka | 120 | 24.39 | (20.8-28.4) |
| Khulna | 59 | 11.99 | (9.4-15.2) |
| Mymensingh | 38 | 7.72 | (5.7-10.4) |
| Rajshahi | 69 | 14.02 | (11.2-17.4) |
| Rangpur | 59 | 11.99 | (9.4-15.2) |
| Sylhet | 30 | 6.1 | (4.3-8.6) |

### Assessment of knowledge and factors associated with knowledge about COVID-19

The average knowledge score for participants was 10.56 (Standard deviation [SD] = 2.86, range 0–14). Among all participants, the range of correct answer rates was between 55.28 and 91.46. About 44.51% of participants were able to provide correct answer for more than 11 questions or obtained scores more than 80%, representing an acceptable level of good knowledge on COVID-19, which was 0.64 more than the average score [Table 2: Respondents’ Knowledge, Attitudes and Practices towards COVID-19.].

**Table 2:** Respondents’ Knowledge, Attitudes and Practices towards COVID-19.

| Questions | Rate of response (%) |  | Mean (SD) | KAP Level (%) |  |
| --- | --- | --- | --- | --- | --- |
| Knowledge about COVID-19 | Yes | No |  | Poor | Good |
| 1. Fever, dry cough and shortness of breath are the main clinical symptoms. | 90.85 | 9.15 | 10.56<br>(2.86) | 55.49 | 44.51 |
| 2. Neck pain/sore throat, tiredness, runny nose, sneezing and diarrhea are fewer common symptoms. | 55.28 | 44.72 |  |  |  |
| 3. Currently there is no effective treatment except symptomatic and supportive treatment. | 71.95 | 8.05 |  |  |  |
| 4. The elder people with chronic illnesses such as diabetic, high BP, heart disease etc. are more likely to be severe cases. | 72.76 | 7.24 |  |  |  |
| 5. Eating or contacting wild animals would result in the infection by the COVID-19 virus. | 60.16 | 39.84 |  |  |  |
| 6. Persons with COVID-2019 without fever can infect others. | 62.4 | 37.6 |  |  |  |
| 7. The COVID-19 virus spreads via respiratory droplets of infected individuals. | 82.11 | 17.89 |  |  |  |
| 8. It is necessary to all to take measures to prevent the infection by the COVID-19 virus. | 56.1 | 43.9 |  |  |  |
| 9. Individuals should avoid going to crowded places such as market, public transportations to prevent the infection. | 83.13 | 16.87 |  |  |  |
| 10. At least 1 meter/ 3 feet is the recommended social distance or physical distance for COVID-19 if go outside of home. | 85.37 | 14.63 |  |  |  |
| 11. Individual should wash hand frequently after coming from outside, before eating or touching mouth, nose, or eyes to prevent the infection. | 91.46 | 8.54 |  |  |  |
| 12. Recommended time for washing hand with soap/ alcohol is minimum 20-30 seconds to prevent the infection. | 78.86 | 21.14 |  |  |  |
| 13. Isolation and supportive treatment are effective ways to reduce the spread of the virus. | 80.28 | 19.72 |  |  |  |
| 14. The immediate observation period is 14 days if anyone contact with someone infected with the COVID-19. | 84.96 | 15.04 |  |  |  |
| **Attitudes towards COVID-19** | Yes | No | Mean (SD) | Poor | Good |
| 1. I agree that COVID-19 will finally be successfully controlled. | 68.5 | 31.5 | 1.24 (0.83) | 51.02 | 48.98 |
| 2. I have confidence that Bangladesh will win the battle against the COVID-19. | 55.28 | 44.72 |  |  |  |
| **Practices towards COVID-19** | Yes | No | Mean (SD) | Poor | Good |
| 1. When I went out, I have avoided crowded place. | 42.62 | 57.38 | 3.17 (1.50) | 76.04 | 23.96 |
| 2. When I went out, I have maintained the recommended social distance of 1 meter or 3 feet. | 63.79 | 36.21 |  |  |  |
| 3. When I went out, I have worn a mask regularly and thoroughly. | 71.31 | 28.69 |  |  |  |
| 4. If I were to go out, I have washed my hand after coming from outside and before eating or touching mouth, nose or eyes regularly and thoroughly. | 76.04 | 23.96 |  |  |  |
| 5. I have maintained the recommended hand washing time of 20-30 seconds regularly and thoroughly. | 63.23 | 36.77 |  |  |  |

Table 3. Association of background characteristics with knowledge towards COVID-19. demonstrates the factors associated with knowledge about COVID-19. Unadjusted model showed that several factors, such as age groups, sex, education, occupation, current living residence, marital status, income level, and administrative regions, were significantly associated with knowledge on COVID-19. The adjusted model, after adjusting for the other variables, showed that education, marital status, family size, monthly income, and administrative regions were significantly associated with knowledge about COVID-19. The higher age groups, such as 46-55, 56-65, and greater or equal to 66, were more likely to have poor knowledge with the lower odds compared to the reference age group below or equal to 25. The female respondents were more likely to have poor knowledge with the lower odds (OR: 0.64; 95% CI: 0.44-0.94) compared to their male counterparts. Among the different occupation groups, farmer (OR:0.42; 95% CI: 0.16-1.09) and day laborer (OR:0.42; 95% CI: 0.2-0.86) were significantly associated with lower knowledge as they had 58% lower odds to have good knowledge compared to reference category, currently not employed. In terms of residence, the rural people (OR:0.44; 95% CI: 0.3-0.64) were significantly lower knowledgeable as they had 56% lower odds to have good knowledge about COVID-19, compared to their urban counterparts. According to administrative regions, the respondents of Rajshahi division (OR:0.25; 95% CI:0.09-0.65) had poor knowledge compared to the respondents from reference regions, Barishal division.

**Table 3.** Association of background characteristics with knowledge towards COVID-19.

| Variables | %(N=492) | Unadjusted Model |  |  | Adjusted Model |  |  |
| --- | --- | --- | --- | --- | --- | --- | --- |
|  |  | OR (SE) | 95% CI | p-Value | OR (SE) | 95% CI | p-Value |
| **Age group** |  |  |  |  |  |  |  |
| ≤ 25 | 17.07 | Ref. |  |  | Ref. |  |  |
| 26-35 | 34.96 | 1.77 (0.48) | (1.05-3.01) | 0.034* | 1.93 (0.73) | (0.92-4.06) | 0.084 |
| 36-45 | 16.87 | 0.57 (0.18) | (0.3-1.05) | 0.072 | 0.79 (0.37) | (0.31-1.99) | 0.616 |
| 46-55 | 11.99 | 0.34 (0.13) | (0.17-0.7) | 0.004* | 0.87 (0.45) | (0.31-2.39) | 0.782 |
| 56-65 | 10.98 | 0.42 (0.16) | (0.2-0.87) | 0.019* | 1.33 (0.73) | (0.45-3.9) | 0.602 |
| ≥ 66 | 8.13 | 0.18 (0.09) | (0.07-0.46) | 0.001* | 0.38 (0.25) | (0.1-1.41) | 0.149 |
| **Sex** |  |  |  |  |  |  |  |
| Male | 65.24 | Ref. |  |  | Ref. |  |  |
| Female | 34.76 | 0.64 (0.12) | (0.44-0.94) | 0.021* | 0.67 (0.2) | (0.38-1.2) | 0.179 |
| **Education Level** |  |  |  |  |  |  |  |
| No education | 15.45 | Ref. |  |  | Ref. |  |  |
| Primary and can sign | 17.68 | 2.38 (0.96) | (1.08-5.25) | 0.031* | 1.88 (0.88) | (0.75-4.69) | 0.175 |
| Secondary and SSC | 20.93 | 2.43 (0.95) | (1.13-5.23) | 0.023* | 1.59 (0.76) | (0.62-4.06) | 0.336 |
| HSC passed or equivalent | 14.23 | 7.88 (3.2) | (3.56-17.45) | 0.001* | 4.58 (2.56) | (1.53-13.72) | 0.006* |
| Higher | 31.71 | 15.53 (5.78) | (7.49-32.2) | 0.001* | 6.07 (3.23) | (2.14-17.23) | 0.001* |
| **Occupation** |  |  |  |  |  |  |  |
| Curr. not employed | 41.06 | Ref. |  |  | Ref. |  |  |
| Service holder | 27.24 | 2.98 (0.69) | (1.89-4.69) | 0.001* | 0.77 (0.41) | (0.27-2.2) | 0.625 |
| Farmer | 5.69 | 0.42 (0.2) | (0.16-1.09) | 0.076 | 0.62 (0.44) | (0.16-2.45) | 0.5 |
| Business man | 11.59 | 1.22 (0.37) | (0.67-2.2) | 0.519 | 0.43 (0.26) | (0.13-1.39) | 0.157 |
| Day laborer | 10.57 | 0.42 (0.15) | (0.2-0.86) | 0.018* | 0.35 (0.21) | (0.11-1.14) | 0.082 |
| Others | 3.86 | 1.73 (0.83) | (0.67-4.45) | 0.255 | 1.15 (0.81) | (0.29-4.6) | 0.845 |
| **Religion** |  |  |  |  |  |  |  |
| Muslim | 92.28 | Ref. |  |  | Ref. |  |  |
| Others | 7.72 | 1.6 (0.54) | (0.82-3.11) | 0.168 | 0.8 (0.41) | (0.29-2.2) | 0.667 |
| **Current living residence** |  |  |  |  |  |  |  |
| Urban | 38.21 | Ref. |  |  | Ref. |  |  |
| Rural | 61.79 | 0.44 (0.08) | (0.3-0.64) | 0.001* | 0.67 (0.18) | (0.39-1.14) | 0.142 |
| **Marital status** |  |  |  |  |  |  |  |
| In a marital relationship | 67.28 | Ref. |  |  | Ref. |  |  |
| Not in a marital relationship | 32.72 | 2.6 (0.51) | (1.76-3.82) | 0.000* | 1.98 (0.55) | (1.14-3.43) | 0.015* |
| **Family size** |  |  |  |  |  |  |  |
| 1-3 members | 22.15 | Ref. |  |  | Ref. |  |  |
| 3-6 members | 64.23 | 0.98 (0.22) | (0.63-1.51) | 0.916 | 2.13 (0.68) | (1.14-3.96) | 0.017* |
| 7 & more | 13.62 | 0.56 (0.18) | (0.3-1.05) | 0.069 | 0.98 (0.46) | (0.39-2.47) | 0.974 |
| **Earning person in family** |  |  |  |  |  |  |  |
| No earning person | 1.22 | Ref. |  |  | Ref. |  |  |
| Single earning person | 51.42 | 0.38 (0.33) | (0.07-2.1) | 0.267 | 0.23 (0.24) | (0.03-1.81) | 0.163 |
| Two & more | 47.36 | 0.42 (0.37) | (0.07-2.32) | 0.318 | 0.23 (0.24) | (0.03-1.82) | 0.163 |
| **Monthly income** |  |  |  |  |  |  |  |
| No earning | 41.06 | Ref. |  |  | Ref. |  |  |
| 1000-10000 | 22.56 | 0.81 (0.2) | (0.5-1.33) | 0.412 | 2.2 (1.11) | (0.82-5.92) | 0.119 |
| 11000-20000 | 16.26 | 1.6 (0.43) | (0.95-2.7) | 0.079 | 1.56 (0.84) | (0.54-4.5) | 0.408 |
| 21000-30000 | 7.93 | 4.5 (1.73) | (2.12-9.56) | 0.001* | 2.51 (1.57) | (0.74-8.54) | 0.139 |
| 31000-40000 | 6.71 | 3.53 (1.4) | (1.62-7.7) | 0.001* | 2.73 (1.75) | (0.78-9.58) | 0.117 |
| >40000 | 5.49 | 10.16 (5.7) | (3.38-30.52) | 0.001* | 14.28 (11.17) | (3.08-66.16) | 0.001* |
| **Availability of running water at home** |  |  |  |  |  |  |  |
| Yes | 91.26 | Ref. |  |  | Ref. |  |  |
| No | 8.74 | 0.51 (0.18) | (0.26-1.01) | 0.052 | 0.54 (0.23) | (0.24-1.24) | 0.146 |
| **Division** |  |  |  |  |  |  |  |
| Barishal | 6.1 | Ref. |  |  | Ref. |  |  |
| Chattogram | 17.68 | 1.4 (0.6) | (0.61-3.23) | 0.429 | 1.38 (0.74) | (0.48-3.94) | 0.548 |
| Dhaka | 24.39 | 0.7 (0.29) | (0.31-1.59) | 0.398 | 1.02 (0.55) | (0.36-2.91) | 0.966 |
| Khulna | 11.99 | 2.37 (1.08) | (0.96-5.81) | 0.06 | 1.58 (0.91) | (0.51-4.89) | 0.426 |
| Mymensingh | 7.72 | 2.01 (0.99) | (0.76-5.3) | 0.161 | 2.87 (1.82) | (0.83-9.94) | 0.096 |
| Rajshahi | 14.02 | 0.25 (0.12) | (0.09-0.65) | 0.005* | 0.22 (0.13) | (0.06-0.73) | 0.013* |
| Rangpur | 11.99 | 0.84 (0.38) | (0.34-2.04) | 0.693 | 3.41 (2.16) | (0.98-11.83) | 0.053 |
| Sylhet | 6.1 | 5.23 (3.07) | (1.66-16.51) | 0.005* | 5.37 (4.01) | (1.24-23.22) | 0.025* |

Fig. 1: Source of knowledge about COVID-19 among participants. shows the sources of knowledge and it indicates television (54%), followed by social media (22%) to be the major sources of knowledge on Covid-19. Other important sources were family members (9%), neighbors (8%), and internet (5%), respectively.

**Fig. 1:**
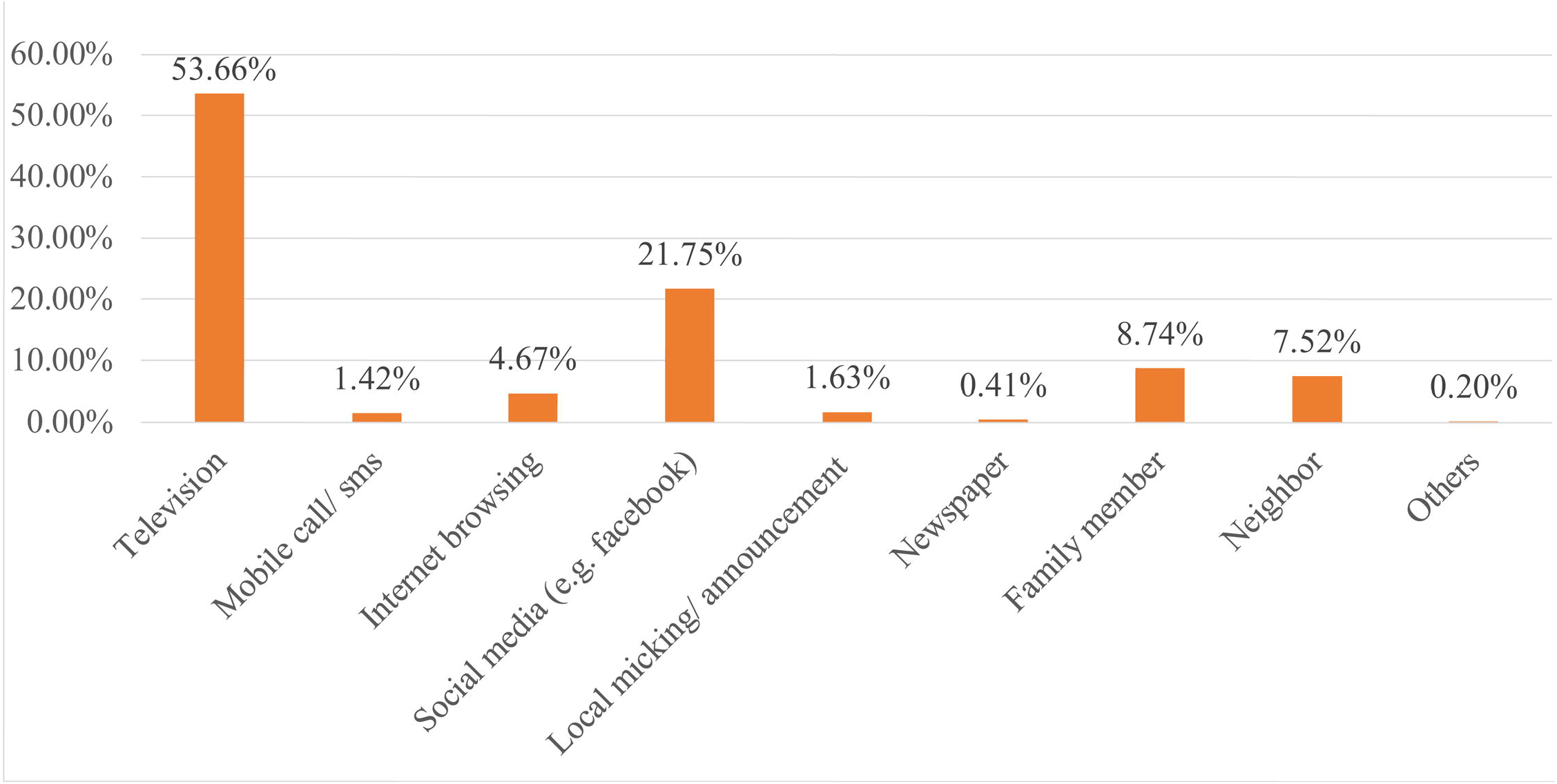
Source of knowledge about COVID-19 among participants.

### Assessment of attitude and factors associated with attitude towards COVID-19

The range of positive attitudes rates for all participants was between 55.28 and 68.50. About 49% of participants were confident and agreed with the 2 questions or obtained scores of 100%, representing an acceptable level of positive attitude towards control and battle against the COVID-19, which was 0.76 more than the average score [Table 2: Respondents’ Knowledge, Attitudes and Practices towards COVID-19.].

Table 4. Association of background characteristics with attitudes towards COVID-19. demonstrates the factors associated with attitudes towards COVID-19. Unadjusted model presented that several factors, such as religion, knowledge about COVID-19, and administrative regions, were significantly associated with attitudes towards COVID-19. Whereas the adjusted model, after adjusting for the other variables, showed that education, occupation, religion, monthly income, knowledge about COVID-18, administrative regions were significantly associated with attitudes towards COVID-19. According to education level, the higher educated people (OR:0.3; 95% CI: 0.12-0.77) were more likely have negative attitudes with the lower odds regarding the controlling of COVID-19 compared to the people with no education. People with income level between 21,000 and 30000, (OR:0.29; 95% CI: 0.09-0.91) and more than 40,0000 (OR:0.25; 95% CI: (0.07-0.9) had higher likelihood of negative attitudes compared to the no income people towards controlling of COVID-19.

**Table 4.** Association of background characteristics with attitudes towards COVID-19.

| Variables | %(N=492) | Unadjusted Model |  |  | Adjusted Model |  |  |
| --- | --- | --- | --- | --- | --- | --- | --- |
|  |  | OR (SE) | 95% CI | p-Value | OR (SE) | 95% CI | p-Value |
| **Age group** |  |  |  |  |  |  |  |
| ≤ 25 | 17.07 | Ref. |  |  | Ref. |  |  |
| 26-35 | 34.96 | 0.98 (0.26) | (0.58-1.64) | 0.927 | 0.95 (0.33) | (0.48-1.89) | 0.89 |
| 36-45 | 16.87 | 0.98 (0.3) | (0.53-1.79) | 0.939 | 1.15 (0.48) | (0.5-2.61) | 0.743 |
| 46-55 | 11.99 | 0.75 (0.26) | (0.38-1.47) | 0.402 | 0.74 (0.33) | (0.3-1.8) | 0.503 |
| 56-65 | 10.98 | 0.66 (0.23) | (0.33-1.31) | 0.231 | 0.66 (0.32) | (0.25-1.72) | 0.394 |
| ≥ 66 | 8.13 | 1.05 (0.41) | (0.5-2.24) | 0.892 | 0.79 (0.42) | (0.28-2.26) | 0.66 |
| **Sex** |  |  |  |  |  |  |  |
| Male | 65.24 | Ref. |  |  | Ref. |  |  |
| Female | 34.76 | 0.87 (0.17) | (0.6-1.27) | 0.476 | 0.9 (0.24) | (0.54-1.5) | 0.684 |
| **Education Level** |  |  |  |  |  |  |  |
| No education | 15.45 | Ref. |  |  | Ref. |  |  |
| Primary and can sign | 17.68 | 0.77 (0.24) | (0.42-1.43) | 0.408 | 0.74 (0.28) | (0.35-1.56) | 0.43 |
| Secondary and SSC | 20.93 | 0.8 (0.24) | (0.44-1.44) | 0.452 | 0.56 (0.22) | (0.26-1.2) | 0.138 |
| HSC passed or equivalent | 14.23 | 1.19 (0.4) | (0.62-2.29) | 0.595 | 0.57 (0.28) | (0.22-1.5) | 0.252 |
| Higher | 31.71 | 0.92 (0.26) | (0.53-1.6) | 0.78 | 0.3 (0.14) | (0.12-0.77) | 0.012 |
| **Occupation** |  |  |  |  |  |  |  |
| Curr. not employed | 41.06 | Ref. |  |  | Ref. |  |  |
| Service holder | 27.24 | 1.16 (0.26) | (0.75-1.8) | 0.505 | 2.2 (1.09) | (0.83-5.81) | 0.111 |
| Farmer | 5.69 | 0.73 (0.3) | (0.33-1.63) | 0.442 | 1.58 (0.99) | (0.46-5.42) | 0.469 |
| Business man | 11.59 | 1.66 (0.51) | (0.92-3.02) | 0.094 | 3.54 (1.96) | (1.2-10.47) | 0.022* |
| Day labor | 10.57 | 1.04 (0.32) | (0.57-1.92) | 0.893 | 2.37 (1.24) | (0.85-6.61) | 0.101 |
| Others | 3.86 | 0.82 (0.4) | (0.32-2.12) | 0.681 | 1.85 (1.28) | (0.48-7.19) | 0.375 |
| **Religion** |  |  |  |  |  |  |  |
| Muslim | 92.28 | Ref. |  |  | Ref. |  |  |
| Others | 7.72 | 5.15 (2.21) | (2.22-11.93) | 0.001* | 3.56 (1.81) | (1.31-9.64) | 0.013* |
| **Current living residence** |  |  |  |  |  |  |  |
| Urban | 38.21 | Ref. |  |  | Ref. |  |  |
| Rural | 61.79 | 0.87 (0.16) | (0.61-1.26) | 0.468 | 0.77 (0.2) | (0.47-1.27) | 0.31 |
| **Marital status** |  |  |  |  |  |  |  |
| In a marital relationship | 67.28 | Ref. |  |  | Ref. |  |  |
| Not in a marital relationship | 32.72 | 1.4 (0.27) | (0.96-2.05) | 0.08 | 1.56 (0.4) | (0.95-2.57) | 0.079 |
| **Family size** |  |  |  |  |  |  |  |
| 1-3 members | 22.15 | Ref. |  |  | Ref. |  |  |
| 3-6 members | 64.23 | 1.32 (0.29) | (0.85-2.05) | 0.214 | 1.49 (0.42) | (0.86-2.61) | 0.157 |
| 7 & more | 13.62 | 1.16 (0.36) | (0.63-2.14) | 0.63 | 1.27 (0.52) | (0.57-2.83) | 0.551 |
| **Earning person in Household** |  |  |  |  |  |  |  |
| No earning person | 1.22 | Ref. |  |  | Ref. |  |  |
| Single earning person | 51.42 | 1.72 (1.51) | (0.31-9.56) | 0.535 | 2.95 (2.8) | (0.46-18.95) | 0.255 |
| Two & more | 47.36 | 2.2 (1.93) | (0.39-12.24) | 0.369 | 4.03 (3.88) | (0.61-26.56) | 0.148 |
| **Monthly income** |  |  |  |  |  |  |  |
| No earning | 41.06 | Ref. |  |  | Ref. |  |  |
| 1000-10000 | 22.56 | 0.74 (0.18) | (0.46-1.18) | 0.199 | 0.42 (0.19) | (0.17-1.03) | 0.057 |
| 11000-20000 | 16.26 | 1.34 (0.36) | (0.79-2.25) | 0.274 | 0.37 (0.19) | (0.14-1.01) | 0.052 |
| 21000-30000 | 7.93 | 1.1 (0.38) | (0.55-2.17) | 0.795 | 0.29 (0.17) | (0.09-0.91) | 0.033 |
| 31000-40000 | 6.71 | 1.11 (0.42) | (0.53-2.31) | 0.79 | 0.32 (0.19) | (0.1-1.06) | 0.062 |
| >40000 | 5.49 | 1.12 (0.46) | (0.5-2.5) | 0.782 | 0.25 (0.16) | (0.07-0.9) | 0.033 |
| **Availability of running water at home** |  |  |  |  |  |  |  |
| Yes | 91.26 | Ref. |  |  | Ref. |  |  |
| No | 8.74 | 0.53 (0.18) | (0.27-1.02) | 0.056 | 0.54 (0.21) | (0.25-1.15) | 0.108 |
| **Knowledge about COVID-19** |  |  |  |  |  |  |  |
| Poor | 55.49 | Ref. |  |  | Ref. |  |  |
| Good | 44.51 | 1.93 (0.35) | (1.34-2.76) | 0.001* | 1.76 (0.45) | (1.07-2.89) | 0.027* |
| **Division** |  |  |  |  |  |  |  |
| Barishal | 6.1 | Ref. |  |  | Ref. |  |  |
| Chattogram | 17.68 | 1.06 (0.46) | (0.45-2.47) | 0.895 | 1.14 (0.53) | (0.46-2.84) | 0.775 |
| Dhaka | 24.39 | 1.11 (0.46) | (0.49-2.51) | 0.804 | 0.86 (0.4) | (0.35-2.12) | 0.741 |
| Khulna | 11.99 | 7.35 (3.74) | (2.71-19.94) | 0.001* | 8.96 (5.04) | (2.98-26.98) | 0.000* |
| Mymensingh | 7.72 | 4.2 (2.2) | (1.5-11.73) | 0.006* | 4.29 (2.47) | (1.39-13.27) | 0.011* |
| Rajshahi | 14.02 | 0.45 (0.21) | (0.18-1.14) | 0.091 | 0.42 (0.22) | (0.15-1.18) | 0.099 |
| Rangpur | 11.99 | 1.35 (0.62) | (0.56-3.3) | 0.504 | 1.54 (0.84) | (0.53-4.49) | 0.426 |
| Sylhet | 6.1 | 3.5 (1.91) | (1.2-10.2) | 0.022* | 1.63 (1) | (0.49-5.42) | 0.425 |

Fig. 2: Distribution of respondents based on went outside of home during lockdown period. shows that about 73% of study population went outside of home during lockdown period. Practices toward COVID-19 were analyzed considering this group. Among the participants, 74% were males who went outside of home.

**Fig. 2:**
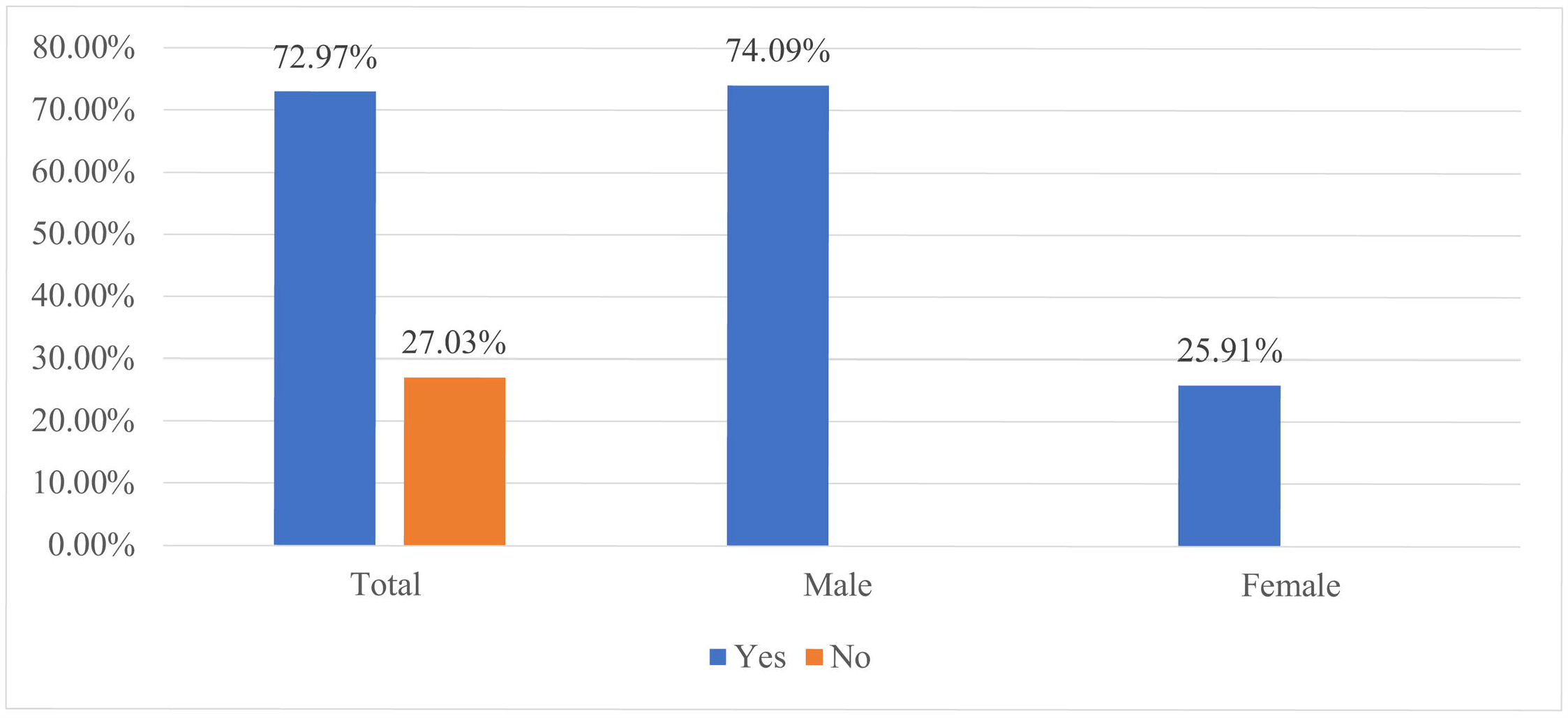
Distribution of respondents based on went outside of home during lockdown period.

Fig. 3. Reasons to go outside of home during lockdown period. shows that about 36% of respondents went outside during lockdown period due to work, followed by 34% to purchases essential goods such as food/ medicine.

**Fig. 3.**
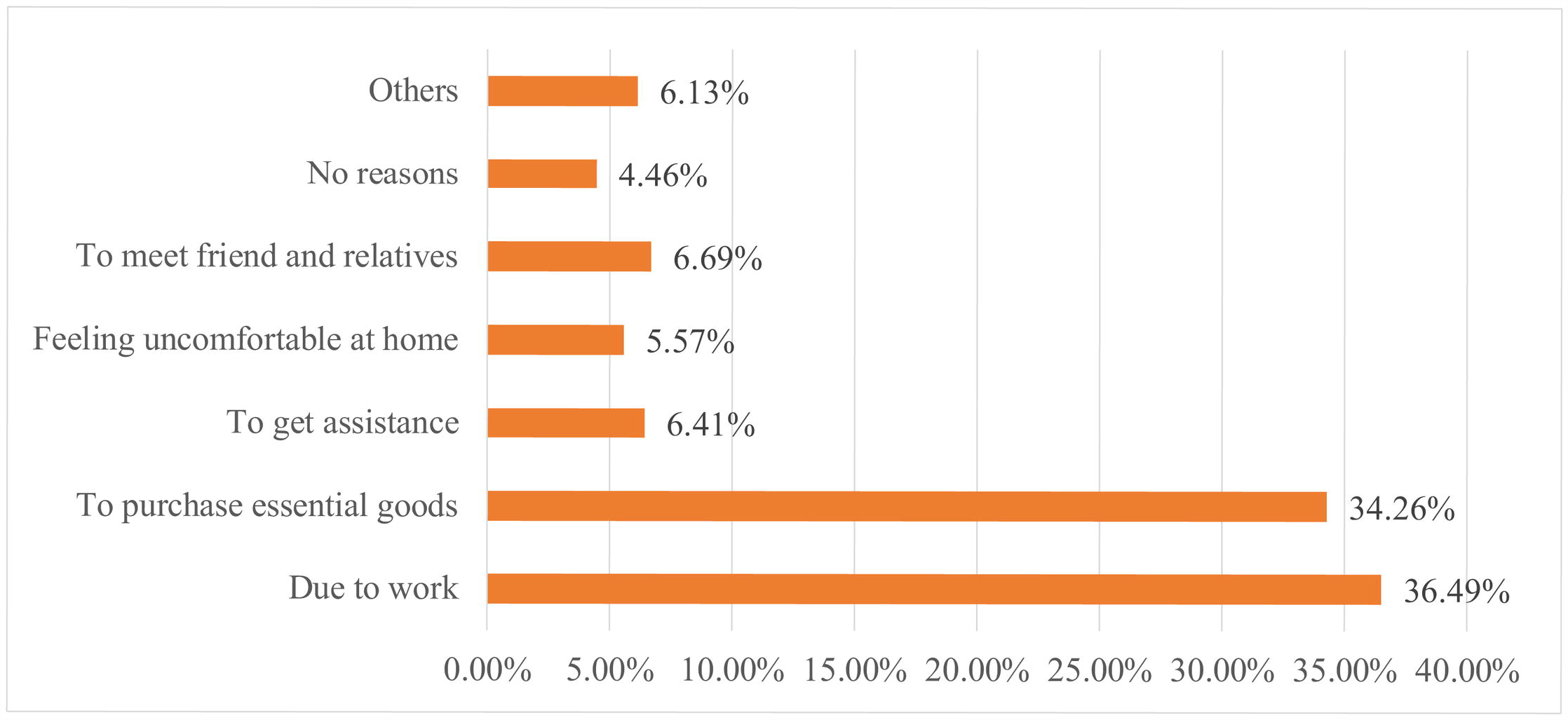
Reasons to go outside of home during lockdown period.

### Assessment of practice and factors associated with practice regarding COVID-19

The average practices score for participants was 3.17 (SD = 1.50, range 0–5). Among all participants, the range of good practices rates was between 42.62% and 76.04%. Overall, about 24% of participants had a favorable practice, and they obtained scores more than 80%, representing an acceptable level of good practice towards COVID-19. This was 0.83 more than the average score [Table 2: Respondents’ Knowledge, Attitudes and Practices towards COVID-19.]. Table 5. Association of background characteristics with practices towards COVID-19. demonstrates the factors associated with practices regarding COVID-19. The unadjusted model showed that several sociodemographic factors, such as age group, education, occupation, residence, income, knowledge and attitude towards COVID-19, and administrative regions were significantly associated with the good practice towards COVID-19. After adjusting for the other variables, the adjusted model showed that religion, number of earning person in family, monthly income, attitudes towards COVID-19, administrative regions were significantly associated with practice towards COVID-19. The results showed that the age group of 46-55 (OR:0.36; 95% CI: 0.13-1.01) had more likelihood of poor practices, compared to the reference age group (age ≤25). The people from other religion category (OR:0.23; 95% CI:0.05-1.01) was found with more likelihood of poor practices, compared to Muslim category. In terms of current residence, rural people (OR: 0.48; 95% CI: 0.29-0.78) had lower practices of safety measures, compared to their urban counterparts. Respondents who from the family with two and more earning persons (OR: 0.06; 95% CI: 0-0.94) were more likelihood to have poor practices, compared to respondents from family with no earning person. According to the income, the respondents with monthly income more than 40,000 (OR:0.08; 95% CI: 0.01-0.67) had more likelihood of poor practices, compared to the their no income reference category.

**Table 5.** Association of background characteristics with practices towards COVID-19.

| Variables | %(N=359) | Unadjusted Model |  |  | Adjusted Model |  |  |
| --- | --- | --- | --- | --- | --- | --- | --- |
|  |  | OR (SE) | 95% CI | p-Value | OR (SE) | 95% CI | p-Value |
| **Age group** |  |  |  |  |  |  |  |
| $\leq 25$ | 16.16 | Ref. | | | Ref. | | |
| 26-35 | 37.33 | 0.92 (0.32) | (0.47-1.82) | 0.81 | 0.49 (0.25) | (0.18-1.33) | 0.16 |
| 36-45 | 19.22 | 0.92 (0.36) | (0.42-1.99) | 0.83 | 0.52 (0.32) | (0.16-1.74) | 0.29 |
| 46-55 | 12.81 | 0.36 (0.19) | (0.13-1.01) | 0.05* | 0.32 (0.24) | (0.07-1.38) | 0.13 |
| 56-65 | 9.47 | 0.42 (0.23) | (0.14-1.26) | 0.12 | 0.52 (0.42) | (0.11-2.52) | 0.42 |
| $\geq 66$ | 5.01 | 0.3 (0.24) | (0.06-1.46) | 0.14 | 0.36 (0.37) | (0.05-2.73) | 0.32 |
| **Sex** |  |  |  |  |  |  |  |
| Male | 74.09 | Ref. |  |  | Ref. |  |  |
| Female | 25.91 | 0.83 (0.24) | (0.47-1.46) | 0.52 | 1.04 (0.47) | (0.43-2.52) | 0.93 |
| **Education Level** |  |  |  |  |  |  |  |
| No education | 12.53 | Ref. |  |  | Ref. |  |  |
| Primary and can sign | 18.94 | 0.99 (0.56) | (0.33-3.01) | 0.99 | 0.69 (0.48) | (0.18-2.69) | 0.59 |
| Secondary and SSC | 20.89 | 1.76 (0.92) | (0.63-4.9) | 0.28 | 1.21 (0.83) | (0.32-4.62) | 0.78 |
| HSC passed or equivalent | 15.32 | 2.67 (1.41) | (0.94-7.53) | 0.06 | 1.13 (0.87) | (0.25-5.12) | 0.87 |
| Higher | 32.31 | 3.29 (1.58) | (1.28-8.44) | 0.01* | 1.51 (1.14) | (0.34-6.63) | 0.58 |
| **Occupation** |  |  |  |  |  |  |  |
| Curr. not employed | 32.03 | Ref. |  |  | Ref. |  |  |
| Service holder | 30.64 | 2.03 (0.63) | (1.11-3.71) | 0.02* | 2.5 (2.13) | (0.47-13.28) | 0.28 |
| Farmer | 6.69 | 0.8 (0.48) | (0.25-2.57) | 0.71 | 6.3 (6.7) | (0.78-50.67) | 0.08 |
| Business man | 15.32 | 1.37 (0.53) | (0.64-2.92) | 0.42 | 2.99 (2.7) | (0.51-17.54) | 0.23 |
| Day labor | 10.86 | 0.73 (0.36) | (0.27-1.94) | 0.53 | 2.51 (2.42) | (0.38-16.6) | 0.34 |
| Others | 4.46 | 0.57 (0.45) | (0.12-2.69) | 0.48 | 0.84 (0.95) | (0.09-7.7) | 0.88 |
| **Religion** |  |  |  |  |  |  |  |
| Muslim | 92.48 | Ref. |  |  | Ref. |  |  |
| Others | 7.52 | 0.53 (0.3) | (0.18-1.58) | 0.25 | 0.23 (0.17) | (0.05-1.01) | 0.05* |
| **Current living residence** |  |  |  |  |  |  |  |
| Urban | 36.49 | Ref. |  |  | Ref. |  |  |
| Rural | 63.51 | 0.48 (0.12) | (0.29-0.78) | 0.001* | 0.49 (0.18) | (0.24-1.02) | 0.06 |
| **Marital status** |  |  |  |  |  |  |  |
| In a marital relationship | 69.64 | Ref. |  |  | Ref. |  |  |
| Not in a marital relationship | 30.36 | 1.41 (0.37) | (0.84-2.35) | 0.19 | 0.95 (0.36) | (0.45-2.02) | 0.89 |
| **Family size** |  |  |  |  |  |  |  |
| 1-3 members | 23.12 | Ref. |  |  | Ref. |  |  |
| 3-6 members | 64.9 | 1.72 (0.55) | (0.92-3.22) | 0.09 | 1.46 (0.63) | (0.63-3.41) | 0.38 |
| 7 & more | 11.98 | 0.88 (0.44) | (0.33-2.36) | 0.80 | 1.4 (0.92) | (0.38-5.1) | 0.61 |
| **No of earning person in family** |  |  |  |  |  |  |  |
| No earning person | 0.84 | Ref. |  |  | Ref. |  |  |
| Single earning person | 49.86 | 0.16 (0.2) | (0.01-1.84) | 0.14 | 0.07 (0.1) | (0-1.22) | 0.07 |
| Two & more | 49.3 | 0.15 (0.18) | (0.01-1.65) | 0.12 | 0.06 (0.08) | (0-0.94) | 0.05* |
| **Monthly income** |  |  |  |  |  |  |  |
| No earning | 32.31 | Ref. |  |  | Ref. |  |  |
| 1000-10000 | 24.79 | 0.51 (0.2) | (0.24-1.11) | 0.09 | 0.38 (0.31) | (0.08-1.9) | 0.24 |
| 11000-20000 | 19.5 | 1.9 (0.64) | (0.98-3.69) | 0.06 | 1.2 (0.94) | (0.26-5.56) | 0.82 |
| 21000-30000 | 9.47 | 2.25 (0.94) | (0.99-5.12) | 0.05 | 0.8 (0.71) | (0.14-4.58) | 0.80 |
| 31000-40000 | 6.96 | 2.86 (1.32) | (1.16-7.07) | 0.02* | 0.86 (0.81) | (0.14-5.46) | 0.88 |
| >40000 | 6.96 | 0.32 (0.24) | (0.07-1.43) | 0.14 | 0.08 (0.09) | (0.01-0.67) | 0.02* |
| **Availability of running water at home** |  |  |  |  |  |  |  |
| Yes | 92.48 | Ref. |  |  | Ref. |  |  |
| No | 7.52 | 1.37 (0.6) | (0.58-3.25) | 0.47 | 1.28 (0.73) | (0.42-3.92) | 0.67 |
| **Knowledge about COVID-19** |  |  |  |  |  |  |  |
| Poor | 54.32 | Ref. |  |  | Ref. |  |  |
| Good | 45.68 | 2.5 (0.64) | (1.52-4.12) | 0.001* | 1.88 (0.71) | (0.91-3.92) | 0.09 |
| **Attitude** |  |  |  |  |  |  |  |
| Negative | 26.74 | Ref. |  |  | Ref. |  |  |
| Positive | 73.26 | 3.77 (1.02) | (2.22-6.4) | 0.001* | 4.47 (1.59) | (2.23-8.98) | 0.001* |
| **Division** |  |  |  |  |  |  |  |
| Barishal | 5.29 | Ref. |  |  | Ref. |  |  |
| Chattogram | 17.27 | 3.15 (2.14) | (0.83-11.97) | 0.09 | 5.65 (4.67) | (1.12-28.53) | 0.04* |
| Dhaka | 23.68 | 1.43 (0.98) | (0.38-5.46) | 0.60 | 2.58 (2.12) | (0.52-12.88) | 0.25 |
| Khulna | 14.76 | 5.97 (4.1) | (1.55-22.95) | 0.01* | 7.61 (6.23) | (1.53-37.84) | 0.01* |
| Mymensingh | 8.08 | 0.62 (0.54) | (0.11-3.43) | 0.58 | 0.48 (0.48) | (0.07-3.4) | 0.47 |
| Rajshahi | 18.94 | 0.33 (0.27) | (0.07-1.64) | 0.18 | 0.79 (0.74) | (0.12-4.96) | 0.80 |
| Rangpur | 7.24 | 0.97 (0.81) | (0.19-4.95) | 0.97 | 2.01 (2.04) | (0.28-14.65) | 0.49 |
| Sylhet | 4.74 | 1.14 (1.02) | (0.2-6.6) | 0.88 | 0.73 (0.78) | (0.09-5.85) | 0.77 |

## Discussion

Our analysis has shown that the knowledge related to Covid-19 of certain socioeconomic groups (e.g., age 46 years of higher, females, those with no education, farmers, day laborers, rural residents, those in a marital relationship, those with a larger family, those with an earning less than BDT 20,000 [USD 236], and residents of Rajshahi division) are significantly lower than the reference category, and most of the people rely on television followed by social media as a source of knowledge. Almost three fourths of the respondents went outside home during the lockdown period and the majority were males (74%), and most went out to purchase essential goods, followed by daily routine work. In terms of practice, rural people lagged behind, as they had 52% lower odds of adhering to appropriate practice measures, compared to their urban counterparts. Finally, we found that a good knowledge and attitude is associated with a better practice of Covid-19 health measures.

The study results showed that higher prevalence of poor knowledge was significantly associated with several demographic and socioeconomic factors. A difference in socioeconomic status contributed to the lower rate of correct COVID-19 knowledge among people in Bangladesh even though the study was conducted after a certain period of the advent of COVID-19 pandemic to Bangladesh. The study observed that aged people tend to have a poor knowledge about COVID-19. This finding is supported by several international studies from developing and developed countries that reported older respondents had poor knowledge on COVID-19 than that of younger [17,20,21]. This fact might be the result of physical condition and loss of cognition status due to ageing associated to watch, read, and understand available and recommended information on COVID-19 considered as barriers to access information about COVID-19 and result in poor knowledge [22]. Familiarity and use of modern technology might be other reasons of poor knowledge among older adults [23,24]. The study observed that farmers and daily laborers were more likely to have poor knowledge about COVID-19. This finding is partially similar with the study in Malaysia and China that the laborers had poorer knowledge [17,26]. Day laborers are one of the major contributors in the informal economy of Bangladesh and depend on their daily wage. Due to the nationwide extended lockdown, they were extremely affected group as they immediately have become jobless. This may indicate limited access to reliable and appropriate information about COVID-19.

Many people did not maintain the lockdown in Bangladesh. Primarily this may be the result of the government’s policies to declare a ‘general holiday’ rather than calling it a lockdown [26,27]. It is worth noting that calling it a holiday rather than a lockdown reduces the gravity of the matter among the public and provide a speculation that people are free to do whatever they want. As a result, many people willingly ignored the stay-at-home or social distancing guidelines and took the opportunity to move to different cities across the country which massively contributed to rapid spread of infection at community level throughout the country [28]. On the hand, the government extended the general holidays without ensuring adequate subsistence support for the poor before lockdown that compelled people to go outside their home [29,30]. Changing the time of lockdown every week might preclude people from taking preparation for the forthcoming days. Moreover, the government’s inability to provide information on how people in lockdown situation can avail essential materials for their life and engage the community groups for meeting essential needs may be the reason of poor practices of safety measures [31,32]. This result reinforces the conclusions of previous studies identifying strict prevention practices and community volunteers mobilization to take care of people under lockdown are the primary solution of reducing spread and control of COVID-19 in China and Vietnam [15,17,33].

Bangladesh is still a predominantly rural based country with only 37% of its population living in urban areas [34]. However, most of the socioeconomic and health indicators are poor for rural areas compared to the urban. For example, 76% of the rural areas are under national electricity grid (urban 92%), 38% of the rural households possess a television (urban 70%), 61% of the rural women got married before age 18 (urban 55%), 60% of the rural women of reproductive age use any contraceptive method (urban 65%), 79% of the rural women received antenatal care from a skilled provider (urban 90%), 45% of the rural women delivered in a health facility (urban 63%), 33% of the rural children under age 5 were stunted (urban 25%), and so on [35]. Similar pattern was observed in our study among the rural people as they had a lower odd of adhering to Covid-19 related hygiene practices. This is particularly troublesome for Bangladesh which has a large number of migrant workers in different countries, returning constantly and spreading out to the rural communities [30,36]. The systematic negligence and ignorance of rural communities towards health policy and programs is observed in several other countries, and this phenomena may pose a higher degree of threat in case of communicable diseases like Covid-19 [20,37,38].

Good knowledge and positive attitudes towards controlling of COVID-19 were associated with the good practices of safety measures. This finding is well recognized in several global studies that a good knowledge and positive attitudes towards COVID-19 leads to improve practices of safety measures [17,20,25,39,40,41]. It is worth mentioning that the consistency of theory-based approaches demonstrates that there is an association among knowledge, belief, and change in human behavior [42]. Adequate and proper knowledge on a specific health emergency is a key modifier of personal belief in changing human behavior [43,44].

Since the level of KAP varies across different socioeconomic groups, we recommend that customized information on Covid-19 should be developed targeting different groups, such as, villagers, slum-dwellers, township residents, urban middle-class, etc. Special emphasis should be given on the groups with lower KAP scores, such as the elderly, females, less educated people, farmers, day laborers, rural residents, those in a marital relationship, those with a larger family, those with a meagre earning, and the residents of Rajshahi division. The information should be clearly and widely circulated through contextually appropriated channels, with emphasis on television and the social media, as these came out to be the major sources of information. Secondly, since many people did not comply with the lockdown directives, the lockdown should be imposed only after ensuring the subsistence support for the poor, arranging emergency requirements of the locked-down community, communicating clearly what to do and not to do during the lockdown period, and clarifying who to consult in case of any unforeseen situation. A voluntary community support group should be engaged in answering to people’s demands. Instead of increasing the duration of lockdown week by week, a tentatively concrete period, in consultation with the epidemiologists, should be imposed on the public so that they can take adequate preparation to stay at home during the instructed period. The term ‘national holiday’ may not convey the right message to the people, so, instead, ‘lockdown’ or any contextually appropriate synonym, in consultation with the communication experts or social scientists, should be used. Special attention should be directed towards the rural communities, where the Covid-19 health practices are found to be the least performed. Finally, since practices are found associated with knowledge and attitude, we recommend that, a scientifically oriented SBCC strategy to be developed in consultation with the relevant experts. To turn these strategies into actions or practices, the religious, cultural, political, and any other community-based forces should be consulted and actively engaged.

### Limitations

The strength of the study is that data were collected from eight administrative divisions throughout the country and participants were surveyed over phone, face to face, and through social media platform from both rural and urban areas. This data collection process improved the generalizability of the findings to the Bangladeshi population. However, this study is not free from limitation. The small sample size of the study may not be representative as compared to the current population in Bangladesh [5]. Another limitation might be the number questions under attitude section where only two questions were considered in the KAP questionnaire to measure the attitude level. The major limitation can be considered with regards to the study design. As a cross-sectional study, causal inferences cannot be drawn here as we cannot assert that the factors which were significantly associated with KAP are certain. Despite these limitations, the findings of the study are believed to motivate and alert policymakers and program implementers who are working on appropriate risk communication and community engagement (RCCE), and SBCC strategies based on the levels of KAP towards COVID-19.

Further research is needed to understand KAP of service providers in Covid-19 pandemic response. Qualitative formative research is useful in designing communication strategies to address the pandemic, and subsequent implementation and evaluation research can generate useful knowledge about the implementation and scaling up of the such strategies in different parts of Bangladesh, and even abroad.

## Conclusions

RCCE is an integral part of pandemic management [45]. In a resource constraint country like Bangladesh, and during a health emergency like Covid-19 pandemic, a study on KAP can render itself to be helpful for the public health decision-makers in designing an evidence-informed and context specific RCCE or SBCC strategies. This study can assist the decisionmakers to identify which groups of people require additional attention for communication. For example, our study identified certain socioeconomic groups with lower level of KAP compared to the reference category. In addition, we figured out the most frequently used source of knowledge, which can be exploited as communication channels which can also be utilized so circulate further knowledge, rules and regulations. The study explored the reasons for nonadherence to lockdown, another important non pharmaceutical intervention against Covid-19, and this information can be supportive to the implementers design a better implementation strategy for lockdown. Finally, this study, by virtue of establishing a positive association between knowledge and attitude with Covid-19 related health practices, highlights the need for an evidence-based informed RCCE and SBCC strategy to foster improved health practices against Covid-19 pandemic.

## Data Availability

Yes - all data are fully available without restriction. All relevant data are within the manuscript and its Supporting Information files.

## Acknowledgement

The authors are thankful to the Public Health Foundation, Bangladesh and its leadership for sponsoring and providing logistic support in conducting the research. We are grateful to Dr. Dipak Kumar Mitra, Professor and Chairman, Department of Public Health, School of Health and Life Sciences, North South University for his valuable inputs in this research. We are also thankful to Mr. Quazi Maksudur Rahmaan, Department of Public Health and Informatics, Jahangirnagar University and Mr. Sourav Paul, Department of Industrial and Production Engineering, Shahjalal University of Science and Technology for their invaluable effort to conduct this study. We would also thanks to the volunteers for their countless support in data collection and a sincere thanks to all respondents participated in the survey.

## Authors’ contribution

**Conceptualization:** Md. Golam Rabbani, Orin Akter, Taufique Joarder

**Data curation:** Md. Golam Rabbani, Orin Akter

**Formal analysis:** Md. Golam Rabbani, Md. Zahid Hasan

**Funding acquisition:** Md. Golam Rabbani, Orin Akter

**Investigation:** Md. Golam Rabbani, Orin Akter, Taufique Joarder

**Methodology:** Md. Golam Rabbani, Orin Akter, Taufique Joarder

**Project administration:** Md. Golam Rabbani, Orin Akter, Taufique Joarder

**Supervision:** Taufique Joarder

**Validation:** Md. Golam Rabbani

**Writing – original draft:** Md. Golam Rabbani, Orin Akter, Md. Zahid Hasan, Nandeeta Samad, Shehrin Shaila Mahmood, Taufique Joarder

**Writing—review and editing:** Md. Golam Rabbani, Orin Akter, Md. Zahid Hasan, Nandeeta Samad, Shehrin Shaila Mahmood, Taufique Joarder

## Supporting information

S1 file. Data sheet.

